# Understanding the roles and experiences of mental health peer support workers in England: a qualitative study

**DOI:** 10.1101/2025.01.16.25320547

**Authors:** Ruth E Cooper, Natasha Lyons, Vicky Nicholls, Una Foye, Prisha Shah, Lizzie Mitchell, Karen Machin, Beverley Chipp, Andrew Grundy, Tamara Pemovska, Nafiso Ahmed, Rebecca Appleton, Julie Repper, Brynmor Lloyd-Evans, Alan Simpson, Sonia Johnson

**Affiliations:** NIHR Policy Research Unit in Mental Health, Institute of Psychiatry, Psychology and Neuroscience, King’s College London, London, UK; NIHR Policy Research Unit in Mental Health, Division of Psychiatry, University College London, UK; NIHR Policy Research Unit in Mental Health (MHPRU) Lived Experience Working Group, Division of Psychiatry, University College London, UK; With-you Consultancy, Merseyside, UK; ImROC (Implementing Recovery through Organisational Change), ImROC Head Office, Nottingham; Florence Nightingale Faculty of Nursing, Midwifery and Palliative Care, Kings College London, UK; North London NHS Foundation Trust, UK

**Keywords:** Peer support, peer workers, qualitative

## Abstract

**Background:** Peer support roles in mental health services are rapidly increasing in the UK and internationally. However, there is wide variation in these roles and limited research exploring the ways in which Peer Support Workers (PSWs) are currently working. We aimed to explore: 1) the distinctive features of PSWs approaches in mental healthcare; 2) the values underpinning the role and 3) the perceived impact of the role.

**Methods:** We conducted semi-structured qualitative interviews with paid mental health PSWs working across a range of settings. We took a co-produced, participatory approach: interviews were carried out by researchers with lived experience of mental health conditions and data were analysed using collaborative methods, guided by general principles of thematic analysis.

**Results:** We interviewed 35 PSWs. Overarching themes identified from iterative analysis included: 1) The centrality of an individualised, flexible, approach, facilitating recovery through sharing lived experiences and building connection. PSWs advocated for service-user needs and most worked in non-clinical ways, offering holistic, recovery-orientated support. Tensions could arise with more clinical approaches. 2) Underpinning values included: i) Recovery is possible: bringing hope, role-modelling and encouraging change, ii) Mutuality: sharing lived experiences to bring empathy and build connection, iii) Person-centred approach: adapting ways of working to the individual, iv) Empowering instead of ‘fixing’ service users. 3) the role had benefits for participants’ own recovery, although its emotional demands could lead to burnout. Participants thought that peer support helped service users feel understood, leading to greater openness and facilitating recovery, although some felt that it may not be right for everyone. Participants felt that PSWs could bring systemic improvements to services and use their lived experience to help teams -meet service user needs.

**Conclusion:** PSWs work in a range of ways, but, a unifying feature is a flexible, person-centred approach, facilitating recovery through shared lived experience. A range of potential benefits of peer work were identified for PSWs and for service users, as well as reports of positive systemic change.

These could be facilitated by recovery-orientated models in services, space for shared learning with PSWs, and flexibility to incorporate PSWs’ unique ways of working.

## Introduction

Peer support work within mental health services refers to employing individuals with personal experience of mental health conditions to provide social, emotional and practical support to others with similar experiences^1^. Peer support promotes recovery through providing hope and an example of recovery to those dealing with mental health difficulties ^2^. The success of peer support is also thought to be through interpersonal connection and the sharing of lived experiences and mental health knowledge ^3,4^. Peer Support may be paid or unpaid ^5,6^, with paid Peer Support Worker (PSW) roles increasingly established in mental health services. Professionalising PSW roles through payment demonstrates the value of the role and rewards work done; it should ensure formal training, supervision and management, and provides a framework for clarifying the remit and boundaries of the role ^7^.

Peer support for people experiencing mental distress is now offered in many countries across the world, leading to the recent development of good practice guidance by the World Health Organization to adapt to the development of peer support services within local contexts. This is to promote the continued expansion of peer support practices in line with recovery and human-rights based approaches, reflecting international efforts to end world-wide coercive practices in mental health. Peer support roles continue to increase rapidly both in the Voluntary, Community, Faith and Social Enterprise (VCFSE) sector and the National Health Service (NHS) in the UK. Peer support is now implemented and recommended across national and international mental health policy guidance, reflecting the perceived value of embedding lived experience support within mental health services ^8–12^.

Wide variation in the roles of PSWs across services has created uncertainty amongst some non-peer staff, service users and peer workers as to what the peer support roles involve ^13,14^. For example, in the NHS roles and titles vary, including PSW, Peer coach, Peer Trainer, and Peer Recovery Worker. Such uncertainty is reflected in the limited literature describing peer support roles, especially regarding the experiences of PSWs themselves, with most literature focusing on barriers and facilitators and impacts of peer support ^13,15–17^. The lack of research exploring peer support roles, and the experiences of peer supporters within these roles, creates challenges for guiding local and national policy and for further research into the effectiveness, experiences and implementation of peer support. For example, we recently conducted one of the largest evidence syntheses of peer support for mental health to date (a systematic umbrella review) ^13^ and found that reviews mainly focused on impacts, barriers and facilitators to peer support rather than the kinds of work that peer workers do in services.

This study aimed to explore the ways in which PSWs are working in mental healthcare in the UK, the values underpinning these roles and their perceived impact. Such understanding is needed to underpin the refinement of approaches to supporting PSWs to use their lived experience effectively to the greatest benefit for people using services. In keeping with key peer support principles, we used methods co-produced with people with lived experience of mental health conditions, including some researchers with experience of providing and receiving peer support, to conduct individual qualitative interviews with paid PSWs in England. We aimed to address the following three research questions:

1. What are the distinctive features of peer support approaches?
2. What values underpin peer support?
3. What is the perceived impact of peer support?

## Methods

This study was conducted by the NIHR Policy Research Unit in Mental Health (MHPRU), which has its main bases at University College London (UCL) and Kings College London and delivers evidence to inform government and NHS policy in England, agreeing a programme of rapid research with policymakers.

We took a coproduced, participatory approach to conducting qualitative interview research ^18,19^. The study is reported according to the COREQ checklist (Consolidated Criteria for Reporting Qualitative Research) (see Supplementary material, S2 Appendix) ^20^. The study was approved by the UCL (University College London) Research Ethics Committee (REF: 19711/001, obtained 9^th^ January 2023).

### Research team and reflexivity

The research team consisted of academic researchers, including some with healthcare professional backgrounds, and lived experience researchers, who have personal experience of using mental health services and/or supporting those who do. Some lived experience researchers also had experience of using or offering peer support or working within the wider peer support sector. A project working group was established which met regularly throughout the project, consisting of the research team and additional experts in peer support (working across both academia and the VCFSE peer support sector). The research team overall was diverse in terms of personal characteristics and perspectives, but academic researchers from minoritized ethnic backgrounds and young people aged twenty-five and under across both Lived Experience and academic researchers were under-represented in our team. Our involvement of people working across a range of roles in the wider field of peer support within the project working group and collaborative approach between lived experience and academic researchers throughout the project ensured researchers with a broad range of perspectives and personal experiences contributed to all research processes.

Interviews were conducted independently by lived experience researchers (KM, BC, PS, LM, VN, NL), with some of these lived experience researchers also having experience working in the field of peer support, including working as peer support worker<u>s</u> in mental health service<u>s</u> and training peer workers. The majority of interviews were facilitated by an academic researcher who was responsible for recording the interviews. Lived experience researchers received training in interviewing skills and qualitative analysis from academic researchers: the majority already had experience of qualitative research. Following interviews, lived experience researchers could debrief with an academic researcher and take part in reflective spaces to discuss any issues arising.

Within peer support there is a range of terminology for peer workers and the people they support. Within this paper we aim to use consistent language where possible, referring to Peer Support Workers (PSWs) and those they support as service users. We acknowledge the range of terms used in the peer support literature and by the participants involved in this study, therefore terminology may sometimes differ to reflect this.

### Participants

Participants were eligible if they were adults (18+ years), who were employed to use their personal experience of mental health conditions in paid PSW roles across a range of mental health services (e.g. NHS, VCFSE). We excluded people: working as volunteer PSWs; working in services solely supporting people who experience organic neurological conditions or substance misuse, or who solely provide peer support through online forums or via online psychoeducation or psychological interventions.

We recruited using purposive sampling to ensure representation of a range of PSW roles, and age groups. Recruitment occurred through circulating the study advert via: i) organisations that provide training for peer support, including organisations who support people who may be underrepresented in peer support research, such as people working with children and young people or with minority ethnic groups; ii) the national and local service user networks known to members of the study team; iii) social media. Interested participants contacted the research team via email, who confirmed eligibility, provided the participant information sheet and offered a phone or video call to discuss the study. Participants gave informed consent to participate in the study. Participants were offered the option of an in person or remote interview by video or telephone.

### Data collection

To develop the topic guide, we held an initial meeting as a research team to discuss potential content. An open document was then shared with all lived experience researchers to add specific topics and questions that would be important to include, guided by experiential expertise, knowledge of peer support practice and current gaps in the literature. We then developed the topic guide through an iterative process of development with regular meetings with the project working group and research team. The topic guide was piloted on the first 10 interviews (See Supplementary material, S1 Appendix). We then held a sense checking workshop with the project working group was to discuss initial findings and agree and make minor changes to the topic guide. The topic guide explored the following areas with PSWs:

- Their roles and unique aspects of their professions.
- The support they provide.
- Their ability to use their lived experience as they would choose to.
- Barriers and facilitators they experience.
- Experiences within mental health teams.
- Supervision, training and career progression.
- Their impact on the mental health system and wider community.

All interviews took place between March and August 2023, remotely via video call on Teams. and were audio-recorded. Before the start of the interview, informed consent was obtained and demographics collected from participants. Participants were informed that their interviewer was a lived experience researcher. Interviews ranged from 30 minutes to 2 hours with the average interview length being 1 hour 14 minutes. Participants were emailed a study debriefing sheet after the interview (giving a list of resources and the option to speak with one of the academic researchers for support, should they feel any distress after the interview). Participants received a gift voucher (£20) for taking part.

### Data analysis

Interviews were recorded, transcribed and anonymised prior to coding. Data were analysed by the research team (lived experience researchers and academic researchers) using co-produced methods based on prior studies ^18,19^ and guided by general principles of thematic analysis ^21^. The analysis process used a coding framework which ensured that insights from all members of the research team, shaped the analysis for this large dataset throughout. The analysis occurred in 5 stages:

1. **Preliminary coding:** the research team (lived experience researchers, n=8, academic researchers, n=2) coded one transcript of an interview from across the dataset, so each researcher coded a different interview. Using a coding matrix, they noted thematic ideas developed and interpreted from the transcript that seemed to articulate core aspects of the interviewee’s experiences and fit within the three research question domains.
2. **Developing a coding framework:** the research team met to review and collate matrices. Similar thematic ideas were grouped together to develop a provisional coding framework. The provisional coding framework was then circulated to the research team and a meeting held to review and refine the framework. A smaller research team (lived experience researchers, n=4, academic researchers, n=2) piloted the framework on 1-2 different interviews each (in NVIVO (version 14), Excel or Word). Another meeting was held to review and refine the framework after piloting, with decisions made about e.g. dividing themes and adding new themes. An initial coding framework was then developed.
3. **Initial coding of data:** the smaller research team coded all interviews using the coding framework (in NVIVO or word).
4. **Refining the coding framework:** throughout interview coding the smaller research team met regularly to discuss whether the interviews they coded offered a good fit with the framework or to agree on revisions to the framework. From this work, novel codes and potential new themes were discussed with the wider team. A revised framework incorporating any adapted or new themes was then produced.
5. **Writing up the analysis:** The revised framework was used by the smaller research team to write up each theme, using example quotes and annotations to write an analytical narrative around the data. To do this, a lived experience researcher paired with an academic researcher to write summaries of the themes, which were organised by each research question. This involved regular meetings to: i) read, review and further consolidate the themes; ii) draft the themes; iii) review and finalise the theme write-up. We made further refinements to the themes through discussion with the wider working group.

## Results

We conducted interviews with 35 eligible participants. Despite confirming eligibility prior to the interview, four participants were excluded post-interview as it emerged during the interview that they did not meet eligibility criteria for their peer support role, e.g. they were working in unpaid roles. No participants withdrew from the study after the interview.

Participant characteristics are shown in Table 1. Most participants were from a white British ethnic background (n=23), were aged between 35-44 years (n=11), and female (n=23). The majority of participants worked in NHS settings (n=21), with fourteen working in other settings such as charities or community-based groups. Considerable variety in job title was found among participants, 11 identified their job as “peer support worker”, and 12 identified as senior peer support roles or peer support coordinator roles. Other job titles included peer consultant, peer coach, peer specialist, peer leader, and peer recovery worker.

**Table 1.** Participant characteristics.

| **Characteristic** | **Category** | **Number** |
| --- | --- | --- |
| Age (years) | 18-24 years | 6 |
|  | 25-34 years | 6 |
|  | 35-44 years | 11 |
|  | 45-54 years | 6 |
|  | 55-64 years | 5 |
|  | >65 years | 1 |
| Gender | Female | 23 |
|  | Male | 10 |
|  | Non-binary/gender self-defined | 2 |
| Ethnicity | White British | 23 |
|  | Asian or South Asian | 5 |
|  | Other white | 3 |
|  | Black Caribbean | 2 |
|  | Black African | 1 |
|  | Any other mixed or multiple ethnic backgrounds | 1 |
| Peer support role | Peer Worker | 23 |
|  | Senior or managerial peer support worker role | 12 |
| Peer support setting | NHS* | 21 |
|  | VCFSE** sector services, e.g. charities, community-based groups, or education settings | 14 |
| Contract type | Permanent contracts | 20 |
|  | Fixed term contracts | 15 |
|  | Self-employed | 1 |
| Employment type | Full time | 17 |
|  | Part time | 18 |
| Length of time in peer role | <1 year | 11 |
|  | 1-2 years | 9 |
|  | 2-5 years | 9 |
|  | 5-10 years | 4 |
|  | >10 years | 2 |

Notes: NHS\* = National Health Service; VCFSE\*\* = Voluntary, Community, Faith and Social Enterprise

We developed 18 themes organised by each of the three research questions below. Themes are summarised in Tables 2, 3 and 4.

1. What are the distinctive features of peer support approaches?
2. What values underpin peer support?
3. What is the perceived impact of peer support?

**Table 2:** What are the distinctive features of peer support approaches? theme summary.

| Theme and description | Illustrative quote |
| --- | --- |
| **Individualised, adaptive and flexible sessions**<br>Peer support sessions were individual or group. Sessions were often open, flexible and unstructured, based on service user needs. Participants would meet service users in a convenient location or in the community, including their home, cafes or mental health services. Sessions encompassed a range of activities and support: emotional and practical support, participants would share their lived experience, discuss strategies and provide resources to manage mental health, or set goals. | *“It’s up to them if they’d like to meet me and we can work out over time possibly some goals that they might like to work towards in peer support. But...they might just want to meet to hear more about my experience, to talk about the experience of psychosis, talk about what happened [in] hospital. If they want to walk their dog, we’ll walk their dog. If they want to go shopping, we’ll go shopping. If they want me to come to their meetings with their psychiatrist or any other meetings- If they want support collecting their prescription or they just want to chat in the living room- It could be any or none of those things.” (PT7, Peer Support Worker)* |
| **Supporting service users holistically: community connections and practical support**<br>Connecting with and supporting service users to go out into the community was an important part | *“We do meet people out in the community. We might be going for coffee. We might also be doing a specific thing. We had peer supporters take clients on dog walks or go to a specific activity together, like going to the gym, or attending an activity where they couldn’t get through the door if they* |
| of the peer worker role. This included, attending community activities with service users (e.g. leisure centres, mother and baby groups) and meeting in cafes. Support would also be provided through providing resources and signposting to other organisations. | *hadn't had someone else with them. Something that we hear a lot in terms of the difference that we've made is, "I wouldn't have been able to make that phone call, send that email, walk through that door without you by my side, without you helping me."* (PT2, Senior Peer Supporter) |
| **Sharing lived experiences with service users to support and bring hope**<br>Participants would share their lived experiences with service users to bring hope and to validate service user experiences. They would provide support based on their experiential knowledge. | *"In terms of my lived experience, I try to share as much as I can and feel comfortable sharing. Obviously, that varies from person to person. Depending on where they're at in their recovery, as well, can really impact how much I share. So, if they've just come out of hospital, I might share my experiences of being in hospital. We talk about differences and similarities there."* (PT9, Peer Support Worker) |
| **Navigating and holding boundaries around disclosure**<br>Boundaries around disclosure and expectations of the peer worker role (e.g. PSWs not acting as therapists), and the PSW role not blurring into friendship with the service user was a key part of the role. For example, sharing enough to build a relationship with the service user but not so much that the PSW would feel vulnerable. | *"It is really easy to lay yourself bare and just be too vulnerable and be sharing your story almost with too many people and in too much detail, that you might reflect on later and think, "Actually, did I overshare? Am I ready for that piece of information to be in somebody else's head, or have I fallen into this place of, 'I really need to prove this thing works,' and so I've been too open?" I have really found that I am constantly evaluating, re-evaluating my own line in the sand where it comes to what I share and what I don't share."* (PT2, Senior Peer Supporter) |
| **Advocating for service user needs**<br>Participants would support service users in interacting with services, e.g. advocating for more or different clinical support. Participants would also draw on their lived experience to facilitate communication between service users and clinicians. | *"within my work, I have to look a lot more at advocacy, and at the issues around the psychiatric system, from a personal perspective, that maybe care coordinators/support workers look at less. So, they're looking more at their role as a professional within the mental health system. Whereas I'm always interrogating my role as a peer, and as a place more in between. ... I'm constantly holding my experiences, and* |
|  | *experiences of friends who I've supported over the years, as the most important aspect, and I'm not forced to go along with a situation because I am a professional and has to agree.* " [PT27, Peer Support Worker] |
| **Cultural inclusiveness**<br>Supporting the specific needs of service users from minority backgrounds was central to some participant's work, e.g. running support groups for people from the same ethnic backgrounds as themselves. | *"What I quite like about my position is that I'm given creative freedom to create and run groups, as long as they are in line with the Trust's policies and protocols. But I quite like that I'm able to create a support service specifically for women of colour, for example. Because as a black woman I find that we're often forgotten and unseen, and I'm definitely not alone in that feeling, so it's nice to support a cause that I feel passionately about."* (PT31, Senior Peer Support Worker) |
| **Working clinically and non-clinically and the challenges between these approaches**<br>Peer workers commonly described working in a non-clinical way, which included not focusing on diagnosis and not trying to 'fix' service users. A smaller number of PSWs who worked in the NHS, worked in more clinical ways, e.g. being involved in discussing medications. There could be challenges between the non-clinical ways of working of peer-workers and more clinical ways of working within teams, especially NHS teams, e.g. peer workers wanted to work with service users for longer than they were able to. | *"For instance, the amount of time I'm spending on someone in a session. I've had someone say, "Well, we need to be sticking to the agreed length of time for sessions so that their expectations don't change and they don't get too attached," and all sorts of stuff. I'm trying to explain, "Well, actually, we're meeting someone on a human level and you don't have to panic that someone is going to get attached if you go 15 minutes over your session time." ...Or if we allow the opportunity for people to come into this building every single day, that's okay too."* (PT2, Senior Peer Supporter) |
| **Working collaboratively with non-peer staff**<br>PSWs would use their lived experience to educate and advise colleagues, reduce stigma and increase compassion towards service users within | *"The other half is speaking with staff members, reminding them when they're speaking, often quite horribly about patients with EUPD that they do have someone in the room that does have those experiences and that maybe it's not best* |
| teams, and help to bring systemic changes to services. Participants could also attend ward rounds and go on joint service user visits with non-peer staff. | *to talk about people in that tone. And to just really hold the space of that this is a service user orientated space, and that we're all sort of working towards the same goals. I think so often there's an us and them mentality, which I tried to sit in between, in my role, which can be challenging, but also is pretty rewarding.*” (PT27, Peer Support Worker) her |

**Table 3:** what values underpin peer support: theme summary.

| Theme | Illustrative quote |
| --- | --- |
| **Recovery is possible**<br>A focus on supporting service users in their recovery underpinned the PSWs approach through being a role model, providing hope and encouraging change in-line with the personal aspirations of those they supported. | *“Clinicians may have worked with service users for a really long time and they’re really, really stuck. Change isn’t being made or there’s a lot of hopelessness, so change feels even harder. Then peer support workers get involved and you can see you start to evoke more hope for that service user through having peer support. I think some of the more [Complex Emotional Needs] patients on that pathway or people who’ve had eating disorders for a really long time and have been quite treatment resistant, suddenly they start to feel more hopeful. Change might start to happen. Not always, but it does, it has in certain cases.” (PT8, Peer Support Worker)* |
| **Mutuality: sharing lived experiences to bring empathy and build connection**<br>Mutuality, building trusting connections with service users, underpinned the work of peer workers. PSWs would empathise with service users by sharing their lived experiences, it was especially helpful where their experiences were similar to service users. These experiences were not only mental health but included being from a particular culture, experiencing poverty or being a veteran. Sharing gave service users | *“There’s a lot of, “Civilians don’t understand this. Civilians don’t understand that.” And one of the good things about the peer support in [service name] is we are all veterans and we [are all lived] experience....So that immediately breaks down two of the biggest barriers, in that I’m a veteran, so I have served, and also I was diagnosed with PTSD.” (PT14, Peer Support Coordinator).* |
| hope and built connection and trust with PSWs. |  |
| **Person-centred and flexible care**<br>A person-centred, flexible approach underpinned peer support sessions, adapting sessions to service user needs. Peer workers were sensitive to how the service user is feeling and what they want from services | *“But I think being flexible also incorporates using your team, using your resources, and if somebody says, “Do you know what, I don't want to do this written piece of work, writing isn't going to work for me, I want to do it in a different way.” “Okay, so let's find a different way. How do you want to do it?” And they may say they want to do it through journaling. They may say they want to do it through artwork. They might say they want to do it through movement. It can be absolutely anything. Vision boarding...” (PT16, Peer Support Worker).* |
| **Empowering instead of ‘fixing’ service users**<br>PSWs aimed to support service users to find their own way through their mental health challenges, instead of trying to ‘fix’ them. | *“I think there is a lot of guilt about not being a good enough mum. ... And I think we can provide emotional support, help people, kind of, come to terms with what's happened to them by just talking about it and realising they're part of a group of women that have had it [postpartum psychosis]... it can empower people and make them feel actually I don't need to feel ashamed of what I went through. I think people often feel a lot of shame and guilt.” (PT18, National Peer Support Service Coordinator)* |

**Table 4:**
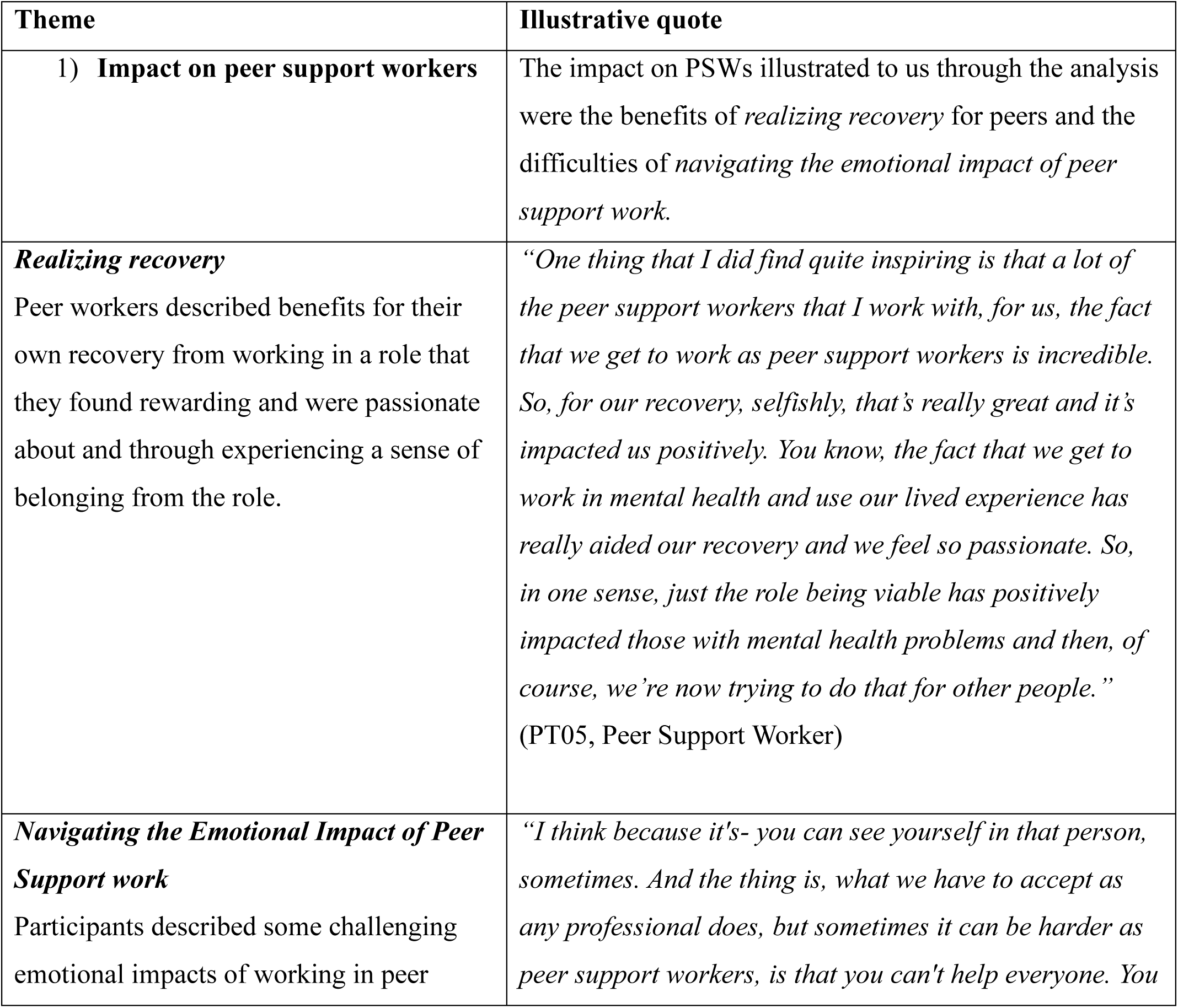
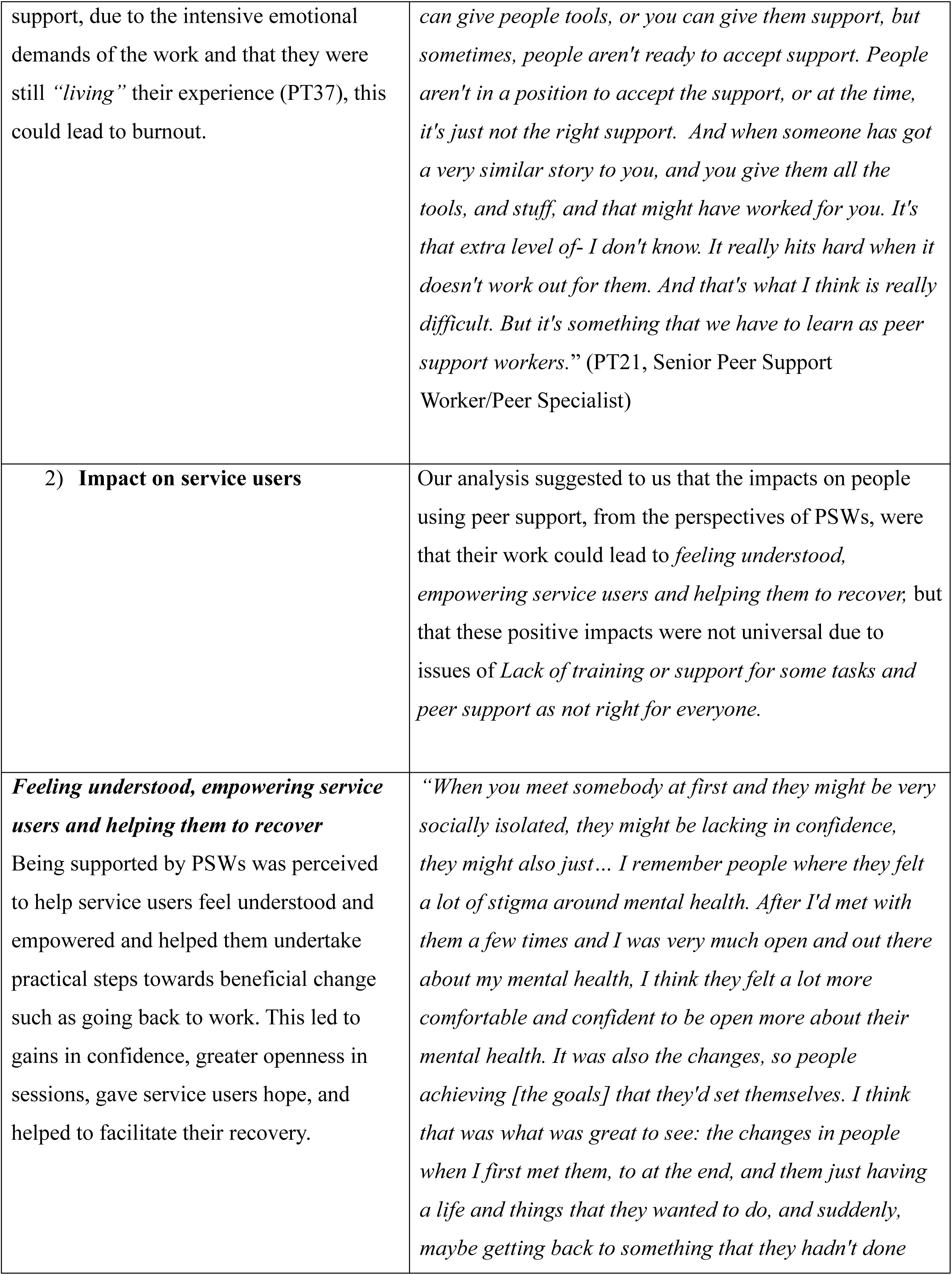
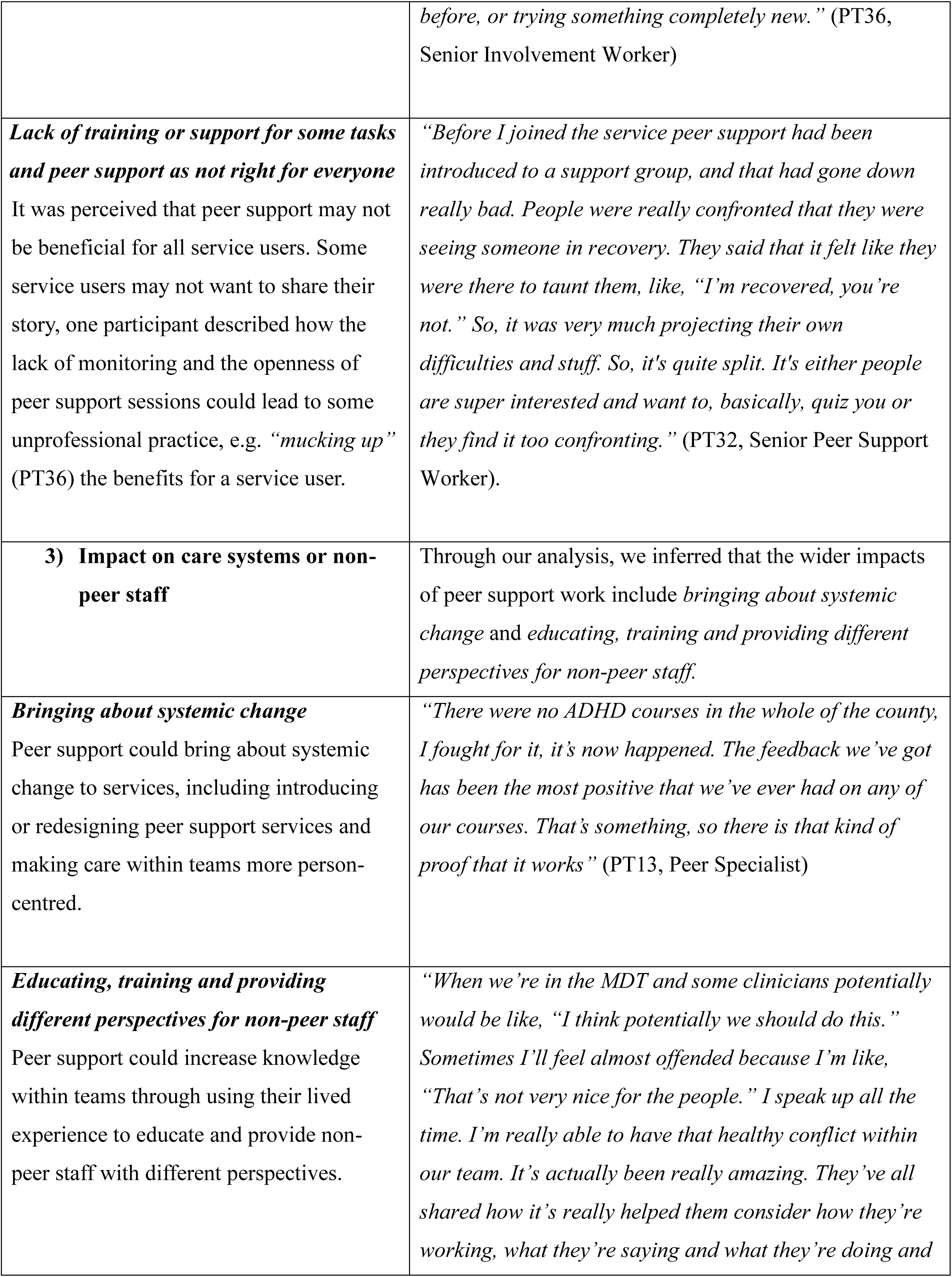
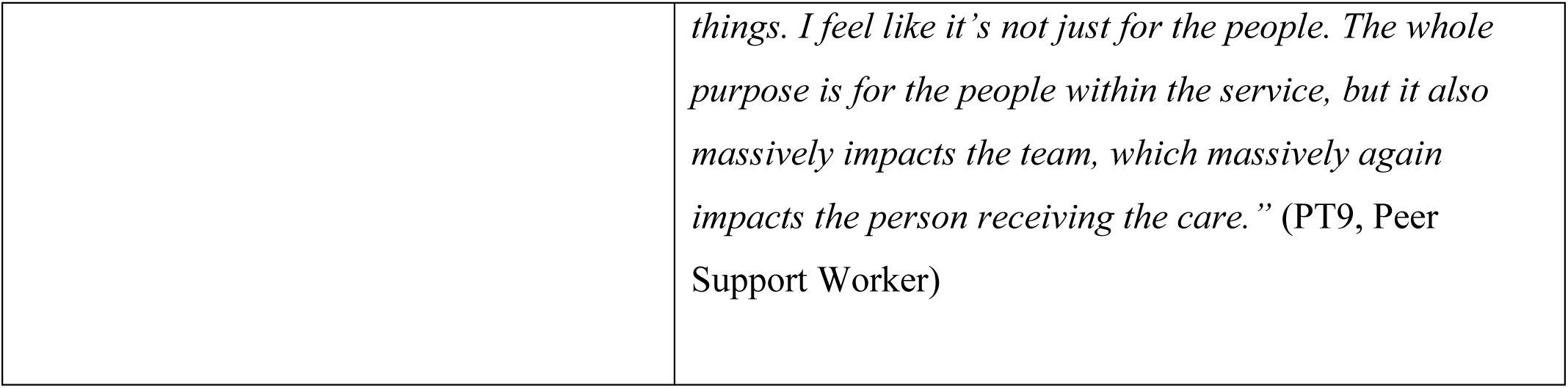
what is the impact of peer support: theme summary.

| Theme | Illustrative quote |
| --- | --- |
| 1) **Impact on peer support workers** | The impact on PSWs illustrated to us through the analysis were the benefits of *realizing recovery* for peers and the difficulties of *navigating the emotional impact of peer support work*. |
| ***Realizing recovery***<br>Peer workers described benefits for their own recovery from working in a role that they found rewarding and were passionate about and through experiencing a sense of belonging from the role. | *“One thing that I did find quite inspiring is that a lot of the peer support workers that I work with, for us, the fact that we get to work as peer support workers is incredible. So, for our recovery, selfishly, that’s really great and it’s impacted us positively. You know, the fact that we get to work in mental health and use our lived experience has really aided our recovery and we feel so passionate. So, in one sense, just the role being viable has positively impacted those with mental health problems and then, of course, we’re now trying to do that for other people.”*<br>(PT05, Peer Support Worker) |
| ***Navigating the Emotional Impact of Peer Support work***<br>Participants described some challenging emotional impacts of working in peer | *“I think because it’s- you can see yourself in that person, sometimes. And the thing is, what we have to accept as any professional does, but sometimes it can be harder as peer support workers, is that you can’t help everyone. You* |
| support, due to the intensive emotional demands of the work and that they were still “*living*” their experience (PT37), this could lead to burnout. | *can give people tools, or you can give them support, but sometimes, people aren't ready to accept support. People aren't in a position to accept the support, or at the time, it's just not the right support. And when someone has got a very similar story to you, and you give them all the tools, and stuff, and that might have worked for you. It's that extra level of- I don't know. It really hits hard when it doesn't work out for them. And that's what I think is really difficult. But it's something that we have to learn as peer support workers.”* (PT21, Senior Peer Support Worker/Peer Specialist) |
| **2) Impact on service users** | Our analysis suggested to us that the impacts on people using peer support, from the perspectives of PSWs, were that their work could lead to *feeling understood, empowering service users and helping them to recover*, but that these positive impacts were not universal due to issues of *Lack of training or support for some tasks and peer support as not right for everyone*. |
| ***Feeling understood, empowering service users and helping them to recover***<br>Being supported by PSWs was perceived to help service users feel understood and empowered and helped them undertake practical steps towards beneficial change such as going back to work. This led to gains in confidence, greater openness in sessions, gave service users hope, and helped to facilitate their recovery. | *“When you meet somebody at first and they might be very socially isolated, they might be lacking in confidence, they might also just... I remember people where they felt a lot of stigma around mental health. After I'd met with them a few times and I was very much open and out there about my mental health, I think they felt a lot more comfortable and confident to be open more about their mental health. It was also the changes, so people achieving [the goals] that they'd set themselves. I think that was what was great to see: the changes in people when I first met them, to at the end, and them just having a life and things that they wanted to do, and suddenly, maybe getting back to something that they hadn't done* |
|  | *before, or trying something completely new.” (PT36, Senior Involvement Worker)* |
| ***Lack of training or support for some tasks and peer support as not right for everyone***<br>It was perceived that peer support may not be beneficial for all service users. Some service users may not want to share their story, one participant described how the lack of monitoring and the openness of peer support sessions could lead to some unprofessional practice, e.g. “*mucking up*” (PT36) the benefits for a service user. | *“Before I joined the service peer support had been introduced to a support group, and that had gone down really bad. People were really confronted that they were seeing someone in recovery. They said that it felt like they were there to taunt them, like, “I’m recovered, you’re not.” So, it was very much projecting their own difficulties and stuff. So, it’s quite split. It’s either people are super interested and want to, basically, quiz you or they find it too confronting.” (PT32, Senior Peer Support Worker).* |
| **3) Impact on care systems or non-peer staff** | Through our analysis, we inferred that the wider impacts of peer support work include *bringing about systemic change and educating, training and providing different perspectives for non-peer staff.* |
| ***Bringing about systemic change***<br>Peer support could bring about systemic change to services, including introducing or redesigning peer support services and making care within teams more person-centred. | *“There were no ADHD courses in the whole of the county, I fought for it, it’s now happened. The feedback we’ve got has been the most positive that we’ve ever had on any of our courses. That’s something, so there is that kind of proof that it works” (PT13, Peer Specialist)* |
| ***Educating, training and providing different perspectives for non-peer staff***<br>Peer support could increase knowledge within teams through using their lived experience to educate and provide non-peer staff with different perspectives. | *“When we’re in the MDT and some clinicians potentially would be like, “I think potentially we should do this.” Sometimes I’ll feel almost offended because I’m like, “That’s not very nice for the people.” I speak up all the time. I’m really able to have that healthy conflict within our team. It’s actually been really amazing. They’ve all shared how it’s really helped them consider how they’re working, what they’re saying and what they’re doing and* |
|  | *things. I feel like it's not just for the people. The whole purpose is for the people within the service, but it also massively impacts the team, which massively again impacts the person receiving the care.” (PT9, Peer Support Worker)* |

## 1. What are the distinctive features of peer support approaches?

An overview of the themes for this research question with illustrative quotes is provided in Table 2. Peer support sessions could be individual, group or a mixture, delivered in person, online (via video call), or over the telephone. Most participants saw service users for a short to medium length of time (e.g. 6-16 sessions) although some participants saw service users longer term (e.g. 1-3 years) or in some cases the support was open-ended. There was often flexibility to increase or decrease the length of time of the support based on service user need. The content of peer support groups varied, including, hearing voices groups, depression and anxiety support groups, and wellbeing groups. Some groups were for people from specific backgrounds, run by PSWs from the same background, such as military veterans and women of colour. A small number of participants offered support to children and young people aged under 25 years old and had used the same service type they worked in themselves.

Participants also described providing support to family members of people using services, such as workshops with carers. Peer support sessions were generally ***individualised, adaptive and flexible***, based on the needs of the service user and were often unstructured, although, some participants described providing structured or unstructured sessions based on service user needs. A small number of participants described offering more structured approaches such as wellness planning, although these were often used flexibly in accordance with the preferences of people using peer support.

Service users were supported ***holistically***, addressing a range of needs through ***connecting with the community*** and being given ***practical support***. Supporting service users to go out into the community could help those who found it difficult to leave their homes and could yield longer term support once the peer worker intervention had finished, through connecting them to community services. Sessions were often held in a location convenient for the service user or in the community, such as at their home or in a café and encompassed a range of activities and support needs. Peer workers would share their lived experience, discuss strategies to manage mental health, provide emotional and practical support and information resources, or set goals, such as leaving the house or getting a job. Peer workers could provide support to attend various activities, such as the leisure centre, the gym, walks, group activities, help with exercising more, or support with starting voluntary work. Participants also viewed peer support groups as communities, bringing people with similar experiences together.

***Sharing lived experiences with service users to support and bring hope*** and validation was central to peer support sessions. Sharing and reflecting on experiences included providing experiential information, such as offering support around self-harm, or giving hope by discussing how they managed to get through difficult mental health experiences. Participants would discuss strategies to manage mental health, e.g. wellness planning, provide emotional and practical support, or set goals. Emotional support was provided through talking, listening and reflecting back to the service user, helping service users feel understood and less isolated. Peer workers were also information sources for service users, including developing and providing resources, e.g., self-harm management. ***Navigating and holding boundaries around disclosure of lived experience*** was key to the peer role. Disclosure to service users was a balance between knowing when sharing was appropriate and sufficient to build a relationship, but not sharing so much that the service user could feel distressed by their story, or that the PSW would feel vulnerable due to oversharing, or that the session did not feel focused on the service user. Boundaries also ensured the relationship with the service user did not blur into friendship and that role expectations were clear, e.g. not acting as therapists to service users.

Participants would also ***advocate for service user needs.*** This included helping them get more or different treatments, such as psychological intervention, obtaining a care assessment, and challenging stigmatising attitudes in mental health teams. Participants would also act as a bridge to facilitate communications between service users and clinicians. Central to the role for some participants was ***cultural inclusiveness***, providing specific support to service users from minority backgrounds. This included running support groups for people from the same ethnic minority background as themselves or helping service users access such services and raising awareness of mental health services in minority communities.

Because the work of peer support workers is guided by their experiential expertise rather than clinical training, their approach was discussed by many participants as being non-clinical and different to usual care. Participants described being a supportive companion in their clients’ recovery journeys through being *“alongside you rather than doing to you”* (PT9), with a strong emphasis on socially focused support. For example, plans for sessions would often be led by clients, and could involve discussing shared experiences or accompanying participants out into the community, such as going for walks, coffee, or to pick up prescriptions or attend appointments. These activities might be steps towards clients’ personal goals that were uniquely identified in peer support sessions, rather than through pre-existing care plans or work with other health professionals within the service. However, some participants who worked in the NHS described taking on more generic clinical roles. This included working in multidisciplinary teams, following clinical guidelines (e.g. CBT guidelines), receiving clinical training, and carrying out clinical duties. Some participants also described using structured scales in their work, such as the DIALOG scale, or using approaches informed by psychological therapies, such as DBT.

A small number of participants, described making recommendations for diagnosis and medication, facilitating or receiving restraint training, focusing on diagnoses, and advising service users about their diabetes.

Within the NHS there could be tensions ***between clinical and non-clinical approaches***. Some PSWs felt that working in the NHS made the PSW role too clinical and professionalised. For example, working more clinically could reduce time spent with service users, e.g. through spending more time completing administrative tasks. Participants described challenges managing service user risk whilst maintaining a trusting relationship, such as having to call emergency services when this might not be what the service user wants, if they are a risk to themselves. Other participants struggled with the length of time they could work with service users, feeling that service users should be able to continue to use services to manage their mental health even if they were perceived to be recovered, or that running over the session time should not be a concern. In some cases, participants could clash with clinical teams in meetings, e.g. where they felt service users needed more empathy.

Participants ***worked collaboratively with non-peer staff.*** They used their experiential knowledge to give advice and provide training to colleagues. Colleagues may ask for support with service users where PSWs ‘experiences aligned, e.g. working with a service user who self-harms, and PSWs could offer more recovery focused ways to manage conditions, even such as ‘safe self-harm’. Participants also attended ward rounds or visited service users jointly with non-peer staff or could ask non-peer staff for support with more complex clients. Some PSWs expressed frustration at stigmatising attitudes towards service users from colleagues and used their lived experience to bring a more compassionate lens to the team, reducing stigma by, for example reminding staff that they have the same diagnosis as the service users that are being talked about negatively. Some participants focused on culture shifts and changes within services, e.g. through being a member of the executive board at the NHS Trust or promoting and ensuring understanding of peer support within services.

## 2. What values underpin peer support?

An overview of the themes for this research question with illustrative quotes is provided in Table 3. The way in which participants worked was underpinned by the distinctive values of peer support work, which were identified in our interviews.

A focus on supporting service users in their recovery and promoting a belief that ***recovery is possible*** underpinned the PSWs’ approach, through being a role model, providing hope and encouraging change. PSWs who had recovered from the same condition were perceived to be ideally positioned to help service users recover, e.g. as role models. In some cases, the peer worker may be one of the first people the service user has spoken to who has experienced the same condition as them and has recovered. Some participants felt that peer workers could give service users more hope than non-peer staff and, through this, bring about change and recovery.

***Mutuality*** underpinned the work of PSWs, participants ***shared their lived experiences to bring empathy and build connection*** with service users, and this was perceived to be a unique aspect of peer support. This was summarised by one participant describing a service user saying, *“I just know you get it”* (PT22). It could be important that PSWs had similar lived experiences to service users not only regarding mental health but also included being from a particular culture, being a migrant or a carer, experiencing poverty, or being a veteran. These shared experiences enabled PSWs to have a holistic understanding of service user’s lives, could normalise service user’ experiences, build trusting connections, provide empathy and hope, and reduce feelings of isolation. Building a strong and trusting interpersonal connection with service users was key to the peer worker role. Participants valued openness and a less hierarchical connection with service users in comparison to clinical roles. This connection enabled service users to be more open about difficult experiences, such as feeling suicidal. Increased trust meant that participants described being able to work with service users whom other staff had been unable to support as either they, or the service-user, had lost trust in the ability of mental health services to help them with their recovery.

***Person-centred approach*** guided the content of peer work sessions, adapting to service user needs for each session. There was a cautious approach to service goals, as opposed to service user’s personal goals and the toolbox of techniques acquired in training, attaching priority to what the service user was wanting from peer support and what they wanted to achieve.

PSWs could have influence and autonomy over how the role was developed and carried out, especially when the PSW service was new, and often used innovative and creative practices.

Participants aimed to ***empower service users instead of trying to ‘fix them’.*** Participants spoke of helping service users find their own ways of moving forward and make their own decisions about what to aim for in working with the PSW. Some spoke of ‘not fixing or trying to change’, but of walking alongside and helping service users to do what they were aiming for, themselves. For example, sharing common or related experience, either by the peer worker or in groups facilitated by the peer worker, could challenge feelings of inadequacy, embarrassment and stigma.

## 3. What is the impact of peer support?

An overview of the themes for this research question with illustrative quotes is provided in Table 4. The themes identified from the analysis indicated impacts of peer support on peer support workers, people using peer support, non-peer staff and healthcare systems.

The diverse and complex work of peer support workers had multiple perceived impacts on participants. These included the positive impact of ***recognising recovery*** in others and benefits for their own recovery journeys, within a role they felt passionate about. Being able to use their past experiences to support others for many made the role *“just so healing in itself”* (PT09). Many peers enjoyed the recognition and positive feedback they received for their work. Other benefits for personal recovery described by participants included experiencing a sense of belonging and feeling more able to be open about their mental health experiences in other areas of their lives. Seeing the benefits of peer support on their clients’ recovery was a highlight of their work, leading to feelings of personal satisfaction, inspiration, and validation among peer workers. For some, the joy they experienced from peer support was in stark contrast to previous roles. Returning to work or finding their ideal profession also brought opportunities for learning more about their own recovery, and that of others, and for developing or broadening a great variety of skills to manage their diverse workloads. Peers described learning from other peer workers, their roles, and from their clients, leading to mutual benefits for peers.

However, the many demands of the role necessitated ***navigating the emotional impact of peer work*.** Most participants were still *“living”* their mental health experiences, so that offering support *“can be tricky when clients bring things that are very close to home”* (PT37). Some participants discussed feelings of powerlessness and frustration when they were unable to give some service users more help, such as when peer support or services more broadly did not work well for an individual. Participants often described working with clients who had experienced trauma and other deeply emotionally affecting experiences. This could lead to emotional burnout and in some cases, peers leaving their posts. The impact of this was greater due to the high levels of responsibility and lack of support experienced by some peer workers. However, other participants felt well supported in their teams and could collaboratively work through distressing experiences. Many participants stressed the need to prioritise self-care to maintain personal wellbeing despite the emotional demands of the role. This included both strategies at work, such as timing appointments to allow space to seek support afterwards and finding time for themselves and for a range of relaxing activities.

Peer workers felt that peer support had a range of benefits for service users. It could help service users ***feel understood, empowered*, *and support their recovery*** and normalise their mental health experiences, reducing feelings of isolation. These benefits were perceived as leading to noticeable changes for service users, including feeling more hopeful and motivated, gaining in confidence, and greater openness in sessions, such as talking more openly about their mental health. Participants felt that their experiential knowledge helped service users to manage their mental health better. Some felt that peer support helped service users who had become disillusioned with mental health services through negative experiences, or through feeling that the services could not help them (as described in our values theme above), to have greater trust in and re-engage with services. Supporting service users to undertake new activities could empower them and provide tools and confidence to continue activities such as going to the gym by themselves. Practical impacts including helping service users leave their homes where they had previously found this difficult. They would help service users into employment, including inspiring and supporting them to become PSWs, help with their housing or financial issues, and support them to socialise more. Advocating for service users could help their recovery, e.g. by helping them access treatments such as psychological therapy.

Some participants described more challenging perceived impacts of peer support for service users. This included PSWs as sometimes ***lack of training or support for some tasks and peer support not being right for everyone.*** One participant described how the lack of monitoring and the openness of session content could lead to unprofessional actions, giving an example of a PSW *“mucking up”* (PT36) the welfare benefits for a service user. Other participants described peer support as not being right for everyone, due to service users not wanting to share their story, the service not being accessed at the right time, e.g. at a time when the service user was not able to engage, or where a relationship between service user and peer worker *“doesn’t work”* (PT10). One participant who worked in an eating disorder service reported some service users finding peer support too confronting, for example when seeing someone who was recovered when they were not. A small number of participants described members of peer support groups upsetting other members e.g. through detailed disclosure of trauma.

Participants felt there were a range of beneficial impacts of peer support for non-peer staff and services. This included ***bringing about systemic change*** in services: for example, PSWs made care within teams more person-centred, helped service users feel more comfortable within services, advocated for service user needs, and introduced new training courses or peer support services, or redesigned these services.

Participants also used their lived experience to ***educate, train and provide different perspectives for non-peer staff***, fostering empathy and closeness within teams. PSWs would enhance the skills of non-peer staff by delivering training or being shadowed by non-peer staff. Participants described using their lived experience to give non-peer staff a different perspective on service users. For example, this could include challenging the way non-peer staff were working with service-users. This led to some non-peer staff re-evaluating their ways of working and noting to participants that this had been helpful to them. Offering different perspectives on people using services also included helping staff support people who experienced specific conditions, such as ADHD, or more complex needs e.g. where the PSW could inform and provide insight based on the complexities of their own experiences, such as of trauma and having multiple conditions. Participants also described mental health teams having better understanding of service user needs through the presence of PSWs, resulting in a better capacity to meet the needs of service users.

## Discussion

### Main findings

We conducted a coproduced qualitative study, interviewing 35 paid PSWs in England to understand the distinctive features of peer support, what values underpin this work, and what impacts it has. Participants worked across a broad range of services and organisations, adopting more traditionally clinical or grass roots approaches to peer support. We found that peers were conducting a wide array of tasks across multiple service types. These could include delivering group and one-to-one support, offering emotional support through shared lived experience, supporting people they worked with to identify and achieve their personal goals for recovery, discussing strategies to cope with experiences of distress based on experiential expertise and co-delivering groups for specific experiences (e.g. anxiety) with non-peer staff. Practical support was also offered within sessions, which included connecting service users with their communities and attending activities with them, often looking outside services for ways to rebuild, further develop or reconnect with their wider lives. The broad and varied nature of their work required the development of diverse skillsets, often at pace.

However, this study has identified unifying and distinctive features of their work, including the flexible nature of the support they offered, involving building a relationship with each individual, taking into account their story and their preferences, and working collaboratively to identify what might help the person most. This enabled them to offer individualised sessions, adapted to the expressed and varying needs of their clients both on that day and throughout their time together, exemplifying an underlying value of a person-centred approach. A unique element of peer work was peers’ use of lived experience to bring hope and validate the experiences of their clients, illustrating an embedded value of mutuality and facilitating recovery through exchanging experiential knowledge. Shared experiences with clients extended beyond mental health to shared culture and a broad array of life experiences, promoting inclusive ways of working.

A non-clinical approach was adopted by most participants, embodying an underlying value of empowering people by walking alongside them rather than trying to “fix” them. However, tensions emerged between the prevailing clinical approaches within many mainstream services and non-clinical ways of working within grass roots models of peer support, with some peers separating and others assimilating clinical ways of working. Peers acted as advocates for those they worked with within these systems, as well as using their experiential expertise to challenge stigma and share their approach with non-peer colleagues, increasing empathic, compassionate and recovery focused ways of working within their wider teams.

Although peers described benefits for their own recovery journeys through supporting those of others, the role had many emotional demands, including being reminded of their own experiences within services or of mental distress. For service users, peer support could help them to feel understood and could normalise their mental health experiences leading to gains in confidence and facilitating recovery. The support also benefitted service users practically, such as helping them go back to work, although some challenging impacts were described, for example, peer support as not being right for everyone, due to some service users not wanting to share their story.

### Findings in the context of the wider literature

The findings of our study build on and update previous investigations of peer support workers’ ways of working across mental health services in England (Gillard et al 2014), adding further detail to these and a wider range of settings.^22^ We have been able to identify some unifying features of peer support approaches across the mental health system. For example, our study encompasses the experiences of peers working within services for people experiencing any form of mental distress to those with specialist provision to people with specific experiences, such as psychosis, complex emotional needs, eating disorders and post-traumatic stress. As highlighted by the work of Gillard and colleagues (2014),^22^ and recent studies,^16^ a distinctive element of the peer role is the unique relationship between peer workers and those they support. Our work further indicates that this unique relationship might rest on their person-centred, adaptive approach of PSWs, and the tailoring of the structure and delivery of support to the unique recovery journey experienced by each service user.

The ways of working of PSWs we identified in our study are aligned with previous peer support literature.^13,16^ The tensions between clinical and non-clinical approaches corresponded with findings from our umbrella review of peer support^13^ and recent literature.^17^ These papers similarly found that challenges to the peer support role included integrating a recovery focused way of working into conventional treatment models^13^ and the dominance of the biomedical model within many mainstream services.^17^ Through bringing together the broad range of service types within our study, we have identified some of the core elements of peer support that unify these distinct approaches within mental health services. Our work highlights the importance of advocacy provided by peers to those they support, offering lived experience perspectives to improve support for people using services, while developing supportive working relationships with many non-peer colleagues and teams.

Many of the values we identified through our analysis closely reflect those suggested to underpin these ways of working within the recent competence framework for peer support workers from Health Education England, such as the concept of non-directive support and respecting the autonomy of service-users within this.^23^ Our study illustrates how these values are embedded in the day-to-day work of peer supporters and presents these alongside the content of this work and the wide-ranging impacts this can have across people and services. The values identified in our study also closely reflect those outlined by many organisations offering or working within peer support, such as those defined by ImROC (Implementing Recovery through Organisational Change).^24^ Our findings support a recently suggested typology of peer support identified by a systematic literature review,^25^ to assist with the implementation and evaluation of these roles across services, which included relationships built on shared lived experience, mutuality and the choice and control of peers’ clients over the support they receive.

Our study indicates that the impacts of the role on many peers highlighted by previous studies^13,15,16^ are shared across a variety of service types. For example, in our recent umbrella review of peer support for the qualitative synthesis of experiences, we found that peer workers experienced improved wellness and recovery from working in the role and increased self-acceptance. However, the role could also negatively impact PSWs wellbeing through reminding them of their mental health condition. For service users, we found peer support gave them hope of recovery, experiential knowledge, normalised their experiences and led to service users feeling empowered.^13^ In a recent mixed methods study of the impact of the peer support role, the role was found to improve PSW wellbeing, including feelings of empowerment and improved self-worth, but could be emotionally demanding, especially early in the role.^15^ Through highlighting the consistency of many of these impacts from individual PSWs to whole teams and services, this indicates the multi-faceted effects of peer support that should be considered with its continued implementation and research into these roles. Through this holistic appraisal of these roles from the perspective of PSWs, our findings illuminate the work needed to shape supportive working environments for PSWs, as these approaches continue to be implemented across an increasing variety of service-types.

### Strengths and limitations

Our inclusion of participants working across the extensive range of peer support roles and services included in this study has highlighted the wide range of ways of working people are working in peer support, as well as identifying its unifying, distinctive features across service contexts. We adopted a collaborative approach between lived experience and academic researchers throughout this study and all interviews were conducted by people with lived experience of mental health conditions. This may have enabled participants to be more open about their perspectives and a deeper exploration of their experiences within their roles ^26,27^. Co-production with lived experience researchers was integral to our research process, ensuring the nuances and complexities of ways of working in peer support were identified through the lens of experiential expertise, and that areas of key importance to people with lived experience were represented from design to final output.

Although the inclusion of a broader range of service types and peer roles than previous studies was a strength of this research, our approach precluded investigation of the distinctive experiences or unique ways of working of PSWs within specific service types, at different levels of seniority or in different sectors, such as NHS and VSCFE settings. The experiences and approaches of senior peers within services remain an under-explored area, which could benefit from further research to unpick these. Given the tensions highlighted between clinical and non-clinical approaches and concerns raised by some participants about protecting the core principles of the role, our realist approach may have precluded more critical interaction with the data. This could have facilitated further exploration of perceptions of tasks, differences between VCSFE and NHS approaches, and alignment between values and ways of working across services. Most peers we interviewed had also been working in their roles for a relatively short time, with 20 participants in post for less than 2 years. This could mean that these participants were still settling into the role, or the role if new may still be subject to development, influencing their responses and possibly not capturing the personal sustainability of performing the roles over a longer period. Two longitudinal analyses that have investigated burnout among peer support workers have found that though there is a small increase in depersonalization^15,28^ and emotional exhaustion^28^ between four^15^ and six months^28^ into the role, compared to when first starting out in it. However, one study found that there was no difference in these measures between six and twelve months into the role,^28^ and the other that changes in burnout were largely unobservable by one year.^15^ Both studies also found that there were no changes in personal achievement across time and that scores on all burnout measures for peer support workers were comparable to other healthcare professional groups, suggesting that the role is likely to be as personally sustainable for many peers as other healthcare workers, at least in the shorter-term.

### Implications for policy and practice

In general, our findings were encouraging for the future of peer support: PSWs across a wide range of settings had a strong sense of the ways in which they could make contributions to service user recovery that were distinctive and helpful, and described considerable satisfaction and personal benefit from their roles. By working in a different way and through their influence on non-peer staff, PSWs can bring vital systemic change to services, and our study provides support for maintaining and continuing to develop such roles. However, we found that some participants felt pressure to conform to more clinical, standardised ways of working or to undertake a range of tasks that were beyond the usual peer roles and for which they may not had access to sufficient training. Pressures on services or confusion about the purpose of PSW roles may contribute to this role confusion. Standardising peer support and adding generic mental health work tasks from the wider team risks losing the flexibility and community-based-based origins of the peer support model and what makes it unique.

Nonetheless, the joy that many participants found from peer support suggests that the broad and varied nature of these the roles may not deter peers from the profession. Moreover, the individualised nature of recovery, day-to-day experiences of mental distress and preferences for support suggests that these roles might necessitate a diversity of tasks, to meet their client-led ways of working. Organisations should ensure a recovery-oriented model of care is operating within services to support the integration of peer workers and sufficient flexibility in approach to service delivery to incorporate the unique ways of working of peer supporters, which may differ from those of clinical staff.

The range of roles and ways of working that are that are compatible with peer support values could usefully be explored further in future research to assist with delineating any boundaries needed around the peer role within services. Practical, socially focused support such as assisting with benefits is an important need but may fall to PSWs by default rather than because it clearly fits with the core purpose of their roles. Teams may need to find other ways of offering such support, such as from specialist benefits advisors, or if a local decision is made that this fits with the role of some PSWs, appropriate training is essential.

Peer work embedded many other elements of good mental health practice, such as culturally inclusive ways of working, and supporting people holistically. We found that peers reported being able to use their experiential expertise to challenge practice and approaches that might negatively impact on people using services. Teams should continue to support opportunities for shared learning and means of adapting practice across services where approaches that optimise the wellbeing of people using services are identified.

### Research implications

Survey based research could be used to investigate the relationships between job characteristics and indicators of psychological and occupational impacts on PSWs, such as wellbeing, job satisfaction and burnout. This may indicate approaches to implementation and support of PSWs that are more tailored to the type of peer support work and service setting.

Similarly, further investigation of the impact of peer work across a variety of service types might uncover any differing impacts of services on the ease of which peers can work in accordance with the values identified here and in previous research^29^ to further tailor service-specific recommendations for implementation. To build on previous research that has investigated the personal impacts of PSWs roles for peers,^15,28^ the longer-term impacts of the role on PSWs beyond one year could be explored in further quantitative and qualitative analysis. This could inform the content of any support or systemic changes needed to assist peer workers pursuing long-term careers in the field.

The broad range of roles that we found peers were working across and the client-led content of their sessions complicates interpretations of the quantitative literature, which has predominantly focused on the effectiveness of peers delivering structured interventions.

Although we did hear from some peers who were using more structured approaches, these were less commonly discussed by participants. This suggests quantitative effectiveness research may often miss the broader picture of peer support work and its impacts on people using peer support and more widely. Our study also highlights the primacy of personal recovery within peer support approaches, and the structuring of the work of many peers around the individual goals, personal priorities and day-to-day needs of those they work with. In-keeping with calls from previous research, this stresses the need for a focus on more personalised^13,30^ and recovery-focused^31^ outcome measures in quantitative evaluations of peer work. To further inform what structured interventions offered by peers might be effective in which contexts, future evaluations of these in specific settings^32^ that clearly specify their content are needed.^32–34^ Examples of these include trials of a peer supported self-management for people leaving crisis services^35^ and a peer supported discharge from inpatient services.^33^

Although many participants assessed the impact of their roles through feedback they had received from both colleagues and clients, further qualitative research is needed to explore the experiences of people receiving peer support and non-peer staff to fully capture the impact of the work of PSWs. There remains a comparable lack of research into the experiences of people using peer support within services, particularly outside of trials. With the expansion of peer support into the vast array of services indicated here, there is an ever-growing need to capture the perspectives of people using services to ensure future developments in peer support continue to be informed by their views.

## Conclusion

Limited research has explored the ways in which Peer Support Workers are currently working. We found PSWs conducted a range of tasks, working flexibly to support the varying needs of service users. However, a unifying feature of their work was their focus on the individual needs of each service user, sharing lived experiences and building a trusting relationship with service users to support recovery. There could be challenges to integrating the PSWs non-clinical ways of working, particularly into NHS services. But a range of beneficial impacts of peer work for PSWs and service users were reported in addition to positive systemic change. Services should ensure space for shared learning with PSWs and clear boundaries around their ways of working. This stresses the need for recovery-oriented models of care in services and flexibility in approach to service delivery to incorporate the unique ways of working of peer supporters.

### Lived experience commentaries

Lizzie Mitchell, Raza Griffiths and Janet Seale, who are members of the MHRPU Lived Experience Researchers Team, wrote individual commentaries on this paper independently of the study.

#### 1) Written by Lizzie Mitchell

As someone who has been a patient with a peer support worker, the finding of them offering holistic, recovery-orientated support echo true. PSW’s acted as my advocate and focused on me as a person – instead of just seeing my illness they saw my personality, hopes, goals and wishes, and the impact recovery would have on my life. My PSW was the first person I had met who had ‘recovered’ and overcome the same difficulties as me, which acted as a turning point and fostered a new sense of hope, self-belief and determination which I’m not sure I would have recovered without.

Accessing mental health services presents huge barriers, and peer approaches focusing on connecting, collaborating, understanding and nurturing individuals can help to mitigate the isolating, lonely and often degrading treatment patients may experience. The clashes between PSW and medical approaches suggest the need for a more recovery-focused, patient-led system, with clinical approaches adopting the collaborative, empowering and adaptive approaches of peers, instead of peers conforming with directive, prescribed clinical approaches.

However, this research only focuses on ‘paid’ peer support workers, missing the uncaptured peers in the charity section, volunteers, and those outside of roles labelled as ‘peer workers’. The ambiguous definition of the role highlight how PSW is often an underutilized, unrecognized profession, relying on the selfless passion and drive of each individual peer worker. As found in the paper, this is not sustainable as peers can end up being burnt out, emotionally drained and underpaid, and move into other roles, and the mutual benefit of the peer role to both patients and staff is lost.

The benefits highlighted in the paper illuminate peer support is a profession to be nurtured, not opposed. The question is how can services collaborate with peers to create a more consistent, sustainable profession, without changing the fundamental beliefs peer work is built on?

#### 2) Written by Raza Griffiths

The Paper highlights the role of Peer Support Workers (PSWs) to establish connection through their own life experiences of distress, thereby fostering hope and recovery, and to strategically challenge stigmatising attitudes and practices in services.

But ultimately, PSWs within the NHS are required to work as gatekeepers, and sign off service users when they meet organisational recovery criteria, irrespective of their wishes. This goes against peer support principles of self-determination, just as having to administer control and restraint, goes against the principle of equal power relationships.

Within statutory services, the PSW role might not be understood or valued by some staff, including HR, because it is a relatively new role, and has values, principles and practices distinct from those of other staff. This can result in scant attention to PSW’s training needs, supervision and safeguarding, leading to ‘burnout’.

Prior to austerity, there were many user-led and community-based peer support groups. These focused on peer support principles of mutuality amongst peers and developing community solidarity. But the focus of many current day PSWs in statutory services is more on individual relationship with service users and less on such communitarian ideas, indicating ideological drift away from original peer support principles

Particularly in the now decimated user led and community sectors, commonality between PSWs and peers was not just in terms of shared experience of mental distress, but in terms of demographic characteristics (like being a Black woman) or life experiences like being a war veteran. Such commonalities could make it easier for peers to identify and feel safe enough to open up.

Those PSWs left in the peer led or community sector face increased challenges as they must constantly meet funding criteria which don’t reflect the wants and aspirations of service users. This is stressful and can disrupt trust and feeling safe in relationships in support groups, which are prerequisites for peer support.

#### 3) Written by Janet Seale

As people with lived experiences of using mental health services or caring for loved ones that do, we welcome any intervention which improves treatment and is recovery focused and the advent of the PSW is all that. Their recovery orientated approach, no diagnosing or prescribing just the powerful tools of lived experience, relating to Service Users as individuals such an approach can be life changing, providing hope and the prospect of recovery. The very existence of the PSW signals that recovery is possible and change achievable. Offering tailored support meeting individual needs moving away from the rigid doctor and patient framework – helping others find their own solutions. The existence of this role reduces stigma offering hope and confidence to a group of people who badly need it.

Although therapeutic the role can be emotionally demanding and may lead to burnout. PSW roles makes services more compassionate, more person and recovery centred and reduces stigma within mental health teams.

The paper captures the distinctive features of the PSW role and the person centred/recovery orientated approach which underpins it. However, the sample size was small only 35 paid PSW, the paper also mentions the lack of standardised training/role description for PSW necessary to prevent any instances of unprofessionalism but at the same emphasises the difficulty of standardising such a unique, individual needs led approach to mental health issues. There are challenges but the benefits to service users, their families and to mental health services that PSW role brings and which this paper demonstrates makes a strong case for expanding and supporting their roles and for more research to be conducted around the standardization of both the role and accompanying job description without any comprise of approach and of the interaction between the PSW role and current mainstream approach of mental health services.

## Supporting information

Consent_document3

consent_document4

## Abbreviations

PSW: Peer Support Worker

VCFSE: Voluntary, Community, Faith and Social Enterprise

## Data Availability

The data that support the findings of this study are available on request from the corresponding author. The data are not publicly available due to privacy or ethical restrictions.

## Acknowledgements

We would like to acknowledge the Mental Health Policy Research Unit team, Lived Experience Working Group members, and Project Working Group who have contributed and supported this work. Special thanks to Raza Griffiths and Janet Seale for their contribution to the paper by providing two independent lived experience commentaries, alongside LM, who provided the third.

## Notes

### Competing Interest Statement

JR is the CEO of ImROC, a provider of peer support training nationally and internationally and is currently one of the organisations commissioned to provide peer support training by NHS England (NHSE). KM is Director of With-You Consultancy, a provider of peer support training nationally and internationally and is currently one of the organisations commissioned to provide peer support training by NHSE.

### Funding Statement

This paper presents independent research commissioned and funded by the National Institute for Health and Care Research (NIHR) Policy Research Programme, conducted by the NIHR Policy Research Unit in Mental Health (MHPRU). The views expressed are those of the authors and not necessarily those of the NIHR, the Department of Health and Social Care or its arm's length bodies, or other government departments. The data that support the findings of this study are available on request from the corresponding author. The data are not publicly available due to privacy or ethical restrictions.

### Author Declarations

The Research Ethics Commitee of University College London (UCL) gave ethical approval for this work (REF: 19711/001, obtained 9th January 2023). All participants provided informed consent prior to enrolment in the study, including consent for publication of anonymised quotes.

## References

1. NHS Health Education England: Peer support workers [Internet]. [cited 2024 Apr 10]. Available from: https://www.hee.nhs.uk/our-work/mental-health/new-roles-mental-health/peer-support-workers

2. Salzer MS, Shear SL. Identifying consumer-provider benefits in evaluations of consumer-delivered services. Psychiatric Rehabilitation Journal. 2002;25(3):281–8.

3. Gillard S. Peer support in mental health services: where is the research taking us, and do we want to go there? Journal of Mental Health. 2019;28(4):341–4.

4. Watson E. The mechanisms underpinning peer support: a literature review. Journal of Mental Health. 2019;28(6):677–88.

5. Galloway A, Pistrang N. “We’re stronger if we work together”: experiences of naturally occurring peer support in an inpatient setting. Journal of Mental Health. 2019;28(4):419–26.

6. National Voices. Peer support: What Is It and Does It Work ? 2015;58.

7. Repper J, Aldridge B, Gilfoyle S, Gillard S, Perkins R, Rennison J. Briefing: Peer Support Workers: Theory and Practice. CentreformentalhealthOrgUk. 2013;1–16.

8. NICE. NHS Long Term Plan » NHS Mental Health Implementation Plan 2019/20 – 2023/24. 2019.

9. Department of Health. The Fifth National Mental Health and Suicide Prevention Plan. The Fifth National Mental Health and Suicide Prevention Plan. 2017.

10. Farmer P, Dyer J. The five year forward view for mental health. The Mental Health Taskforce. 2016.

11. Integrated personal commisioning. Community capacity and peer support: summary guide. NHS England. 2017;66.

12. Mental Health Commission of Canada: Peer Support [Internet]. Available from: https://mentalhealthcommission.ca/what-we-do/access/peer-support/

13. Cooper RE, Saunders KRK, Greenburgh A, Shah P, Appleton R, Machin K, et al. The effectiveness, implementation, and experiences of peer support approaches for mental health: a systematic umbrella review. BMC Medicine. 2024;22(1).

14. Collins R, Firth L, Shakespeare T. “Very much evolving”: a qualitative study of the views of psychiatrists about peer support workers. Journal of Mental Health. 2016;25(3):278–83.

15. Gillard S, Foster R, White S, Barlow S, Bhattacharya R, Binfield P, et al. The impact of working as a peer worker in mental health services: a longitudinal mixed methods study. BMC Psychiatry. 2022;22(1):1–18.

16. Tisdale C, Snowdon N, Allan J, Hides L, Williams P, de Andrade D. Youth Mental Health Peer Support Work: A Qualitative Study Exploring the Impacts and Challenges of Operating in a Peer Support Role. Adolescents. 2021;1(4):400–11.

17. Chisholm J, Petrakis M. Peer Worker Perspectives on Barriers and Facilitators: Implementation of Recovery-Oriented Practice in a Public Mental Health Service. Journal of Evidence-Based Social Work (United States). 2023;20(1):84–97.

18. Gillard S, Dare C, Hardy J, Nyikavaranda P, Rowan Olive R, Shah P, et al. Experiences of living with mental health problems during the COVID-19 pandemic in the UK: a coproduced, participatory qualitative interview study. Social Psychiatry and Psychiatric Epidemiology. 2021;56(8):1447–57.

19. Shah P, Hardy J, Birken M, Foye U, Rowan Olive R, Nyikavaranda P, et al. What has changed in the experiences of people with mental health problems during the COVID-19 pandemic: a coproduced, qualitative interview study. Social Psychiatry and Psychiatric Epidemiology. 2022;57(6):1291–303.

20. Tong A, Sainsbury P, Craig J. Consolidated criteria for reporting qualitative research (COREQ): a. Int J Qual Health Care. 2007;19(6):349–57.

21. Braun V, Clarke V. Using thematic analysis in psychology. Qualitative Research in Psychology. 2006;3(2):77–101.

22. Gillard S, Edwards C, Gibson S, Holley J, Owen K. New ways of working in mental health services: a qualitative, comparative case study assessing and informing the emergence of new peer worker roles in mental health services in England [Internet]. Southampton (UK): NIHR Journals Library; 2014 [cited 2025 Jan 8]. (Health Services and Delivery Research). Available from: http://www.ncbi.nlm.nih.gov/books/NBK373837/

23. Health Education England. The Competence Framework for Mental Health Peer Support Workers. Part 2: Full listing of the competences [Internet]. 2020 [cited 2025 Jan 8]. Available from: https://www.hee.nhs.uk/our-work/mental-health/new-roles-mental-health/peer-support-workers

24. ImROC: the values that underpin peer support work. [Internet]. [cited 2025 Jan 8]. Available from: https://imroc.org/peer-support-old/the-values-that-underpin-peer-support-work/

25. Kotera Y, Newby C, Charles A, Ng F, Watson E, Davidson L, et al. Typology of Mental Health Peer Support Work Components: Systematised Review and Expert Consultation. Int J Ment Health Addiction [Internet]. 2023 Aug 14 [cited 2023 Aug 29]; Available from: https://link.springer.com/10.1007/s11469-023-01126-7

26. Faulkner A. Principles and motives for service user involvement in mental health research. In: Amering M, Schrank B, Wallcraft J, Wiley I, editors. Handbook of service user involvement in mental health research. In Chichester: Wiley-Blackwell; 2009. p. 13–24.

27. Gillard S, Borschmann R, Turner K, Goodrich-Purnell N, Lovell K, Chambers M. ‘What difference does it make?’ Finding evidence of the impact of mental health service user researchers on research into the experiences of detained psychiatric patients. Health Expectations. 2010;13(2):185–94.

28. Park SG, Chang BH, Mueller L, Resnick SG, Eisen SV. Predictors of Employment Burnout Among VHA Peer Support Specialists. PS. 2016 Oct;67(10):1109–15.

29. Gillard S, Foster R, Gibson S, Goldsmith L, Marks J, White S. Describing a principles-based approach to developing and evaluating peer worker roles as peer support moves into mainstream mental health services. Mental Health and Social Inclusion. 2017 Jun 12;21(3):133–43.

30. Law D. The goal-based outcome (GBO) tool: guidance notes [Internet]. London, UK: MindMonkey Associates; 2019 [cited 2025 Jan 8]. Available from: https://www.goals-in-therapy.com/

31. Chinman M, George P, Dougherty RH, Daniels AS, Ghose SS, Swift A, et al. Peer Support Services for Individuals With Serious Mental Illnesses: Assessing the Evidence. PS. 2014 Apr;65(4):429–41.

32. Lyons N, Cooper C, Lloyd-Evans B. A systematic review and meta-analysis of group peer support interventions for people experiencing mental health conditions. BMC Psychiatry. 2021 Jun 23;21(1):315.

33. Gillard S, Bremner S, Patel A, Goldsmith L, Marks J, Foster R, et al. Peer support for discharge from inpatient mental health care versus care as usual in England (ENRICH): a parallel, two-group, individually randomised controlled trial. Lancet Psychiatry. 2022 Feb;9(2):125–36.

34. White S, Foster R, Marks J, Morshead R, Goldsmith L, Barlow S, et al. The effectiveness of one-to-one peer support in mental health services: a systematic review and meta-analysis. BMC Psychiatry. 2020 Nov 11;20:534.

35. Johnson S, Lamb D, Marston L, Osborn D, Mason O, Henderson C, et al. Peer-supported self-management for people discharged from a mental health crisis team: a randomised controlled trial. The Lancet. 2018 Aug 4;392(10145):409–18.

