## Supplementary material for "Understanding the roles and experiences of mental health peer support workers in England: a qualitative study": Consent_document3

Consent for Publication

I confirm that I give my consent for the publication of my Lived Experience Commentary in the manuscript titled:

I confirm that I give my consent to be named as the author of the Commentary on the manuscript.

Signed: Janet Seale

JANET SEALE

………………………………………………..

(please add your name above)
